# Computational analysis of congenital heart disease associated SNPs: Unveiling their impact on the gene regulatory system

**DOI:** 10.1101/2024.03.20.24304537

**Authors:** Shikha Vashisht, Costantino Parisi, Cecilia L. Winata

## Abstract

Congenital heart disease (CHD) is a prevalent condition characterized by defective heart development, causing premature death and stillbirths among infants. Genome-wide association studies (GWASs) have provided insights into the role of genetic variants in CHD pathogenesis through the identification of a comprehensive set of single-nucleotide polymorphisms (SNPs). Notably, 90-95% of these variants reside in the noncoding genome, complicating the understanding of their underlying mechanisms. Here, we developed a systematic computational pipeline for the identification and analysis of CHD-associated SNPs spanning both coding and noncoding regions of the genome. Initially, we curated a thorough dataset of SNPs from GWAS-catalog and ClinVar database and filtered them based on CHD-related traits. Subsequently, these CHD-SNPs were annotated and categorized into noncoding and coding regions based on their location. To study the functional implications of noncoding CHD-SNPs, we cross-validated them with enhancer-specific histone modification marks from developing human heart across 9 Carnegie stages and identified potential cardiac enhancers. This approach led to the identification of 2,056 CHD-associated putative enhancers (CHD-enhancers), 38.9% of them overlapping with known enhancers catalogued in human enhancer disease database. We identified heart-related transcription factor binding sites within these CHD-enhancers, offering insights into the impact of SNPs on TF binding. Conservation analysis further revealed that many of these CHD-enhancers were highly conserved across vertebrates, suggesting their evolutionary significance. Utilizing heart-specific expression quantitative trait loci data, we further identified a subset of 63 CHD-SNPs with regulatory potential distributed across various cardiac tissues. Concurrently, coding CHD-SNPs were represented as a protein interaction network and its subsequent binding energy analysis focused on a pair of proteins within this network, pinpointed a deleterious coding CHD-SNP, *rs770030288*, located in C2 domain of *MYBPC3* protein. Overall, our findings demonstrate that SNPs have the potential to disrupt gene regulatory systems, either by affecting enhancer sequences or modulating protein-protein interactions, which can lead to abnormal developmental processes contributing to CHD pathogenesis.

**Authors Summary:** Congenital heart disease (CHD) is a common condition with defects in heart development present from birth. CHD symptoms can range from mild to severe, often requiring early intervention or surgery. Over the years, numerous research studies have indicated the association of single nucleotide polymorphisms (SNPs) with CHD. However, the challenge arises from the fact that the majority of these variants are located within the noncoding portion of the genome, making it difficult to comprehend their mechanism of action. Here, we present a systematic computational pipeline to identify SNPs associated with CHD, in both protein-coding and noncoding regulatory elements – specifically, enhancers. Utilizing this pipeline, we established a collection of putative enhancers containing CHD-SNPs. Within these enhancers, several transcription factor binding sites (TFBSs) related to heart developmental processes were identified. The presence of SNPs in these sites may potentially impact the binding of TFs necessary for the expression of genes targeted by these enhancers. Additionally, some of these enhancers were also found to be evolutionary conserved, suggesting their functional relevance. Concurrently, we identified coding variants which can alter the protein-protein interactions in a protein interaction network. Taken together, our study provided critical insights into the role of genetic variants in the pathological mechanism of complex human diseases, including CHD.

## Introduction

Congenital heart disease (CHD) is a defect of the heart caused by abnormalities in the process of its development. It is one of the most common birth defects characterized by a wide range of structural deformities of the heart and great vessels, affecting 10 per 1000 (∼1%) neonates worldwide, along with 10% of stillbirths (1–4). The complex multifactorial etiology of CHD presents a major challenge towards understanding its clear pathological mechanism. A large number of protein-coding genes are known to be implicated in CHD (5–7). However, whole genome sequencing (WGS) data from thousands of CHD patients and genome-wide association studies (GWAS), have revealed that the majority (∼95%) of CHD-causing genetic variants fall within the noncoding fraction of the genome (8,9). Despite tremendous progress made towards their identification, we still lack clarity on how noncoding genetic variants associated with CHD-traits are implicated in defective heart formation (10). Hence, there is a fundamental need to discover the contribution of noncoding variants towards CHD development and its manifestation to offer novel insights into the genetic basis of CHD.

Noncoding genetic variants could alter normal developmental processes by disrupting the function of enhancers (11). It is estimated that the number of disease-associated variants impacting enhancer function far exceeds that affecting protein-coding genes (12,13). Enhancers are a major type of *cis*-regulatory element in the genome which regulates the expression of downstream target genes. They serve as binding sites for transcription factors (TFs) which are necessary for the activation of target gene expression. Genetic variants within enhancers could therefore disrupt TF binding sites (TFBSs), causing the failure of TF binding and ultimately loss of enhancer function associated with disease (14,15). Despite the wealth of data available on putative enhancers, there is still a lack of systematic analyses focused on particular biological context within development and disease. Moreover, validating the enhancer activity of these millions of regions, as well as ascribing their function and target genes still poses an uphill task. Further, the ability to link enhancers to their putative target genes could provide additional critical insights into their possible role in specific biological processes, particularly if the function of the target is already known. A comprehensive map of genetic interactions consisting of not only protein coding genes, but also enhancers regulating their expression, could therefore contribute valuable insight into their function and mechanism in regulating specific biological processes.

In this study, we present a catalogue of 2,056 putative human cardiac enhancers and a protein-protein interaction network associated with CHD through a systematic compilation and analyses of CHD-specific GWAS data. To achieve this, we devised a computational pipeline to process SNPs spanning both noncoding and coding regions of the genome. Using noncoding CHD-SNPs data, we predicted CHD-enhancers through a rational integration of epigenomics data of human heart organogenesis followed by conservation analyses and intersection with expression quantitative trait loci (eQTL) data to establish their regulatory potential. The identified set of cardiac enhancers contained specific TF binding sites specifically linked to heart function which further support their functional relevance.

Parallel investigation into coding CHD-SNPs focused on their potential impact on protein-protein interaction stability, we first constructed a protein-protein interaction network (PPIN, **Fig S1**) and analyzed the interaction between two crucial cardiac sarcomeric proteins—cardiac myosin binding protein-C (*MYBPC3*) and cardiac α-actin (*ACTC1*) (16). Prior studies revealed a substantial number of individuals with hypertrophic cardiomyopathy (HCM) and dilated cardiomyopathy exhibit alterations in sarcomeric proteins (17,18). Examining changes in binding energy within the three-dimensional structures of these proteins revealed a deleterious missense SNP (c.1256G>A; p.Arg419His; *rs770030288*). This mutation exhibited a notably higher potential to destabilize the interaction between these proteins, emphasizing the significance of this mutation in the context of protein stability and cardiac function. Taken together, these results highlight the potential significance of our approach in contributing to the understanding of genetic factors influencing CHD, providing a holistic perspective on both regulatory and coding elements.

## RESULTS

### 1. A compilation of CHD-associated single nucleotide polymorphisms and their annotations

To identify CHD-associated SNPs, we retrieved data from single nucleotide polymorphism (SNP) resources based on Genome-wide Association Studies (GWAS), including NHGRI-EBI GWAS-catalog (19) and ClinVar (20,21) databases. This systematic approach resulted in a cumulative set of 15,876 CHD-SNPs (computational pipeline illustrated in **Fig 1A**). As of August, 2023, the GWAS-Catalog contains a total of 5,39,949 SNPs while ClinVar database consists of a total of 4,609,367 variations including SNPs and other types of genomic variations. In GWAS-catalog, out of 5,39,949 SNPs, 1,884 unique statistically significant (p-value < 10e-6) SNPs were found to be associated with CHD-related traits (4,22–25) (CHD-SNPs; **Table S1 and S2**). Consequently, ClinVar consisted of 4,609,367 variations, out of which we retrieved 15,248 unique CHD-specific variations which consisted of 13,992 CHD-SNPs and other types of variations, including 629 deletions, 287 duplications, 107 indels, 43 insertions and 190 microsatellites (**Table S3**). Further, ANNOVAR (26) annotation of CHD-related variants revealed that, from GWAS-catalog, 33.55 % and 43.63% lies within the intergenic and intronic regions of the genome while only 4.46% of the variants were from the exonic part of the genome (**Fig 1B**). On contrary, ClinVar database consisted of a large percentage (83.54%) of CHD-SNPs within the exonic part of the genome while only 0.01% and 7.03% of the CHD-SNPs were present on intergenic and intronic regions, respectively (**Fig 1C**). These differences exist mainly due to the fact that more than 80% of ClinVar associations result from whole exome sequencing studies (WES), thus predominantly capturing the protein-coding part of the genome. Additionally, ClinVar primarily deposits fully curated and clinically tested genetic variations.

**Fig 1:**
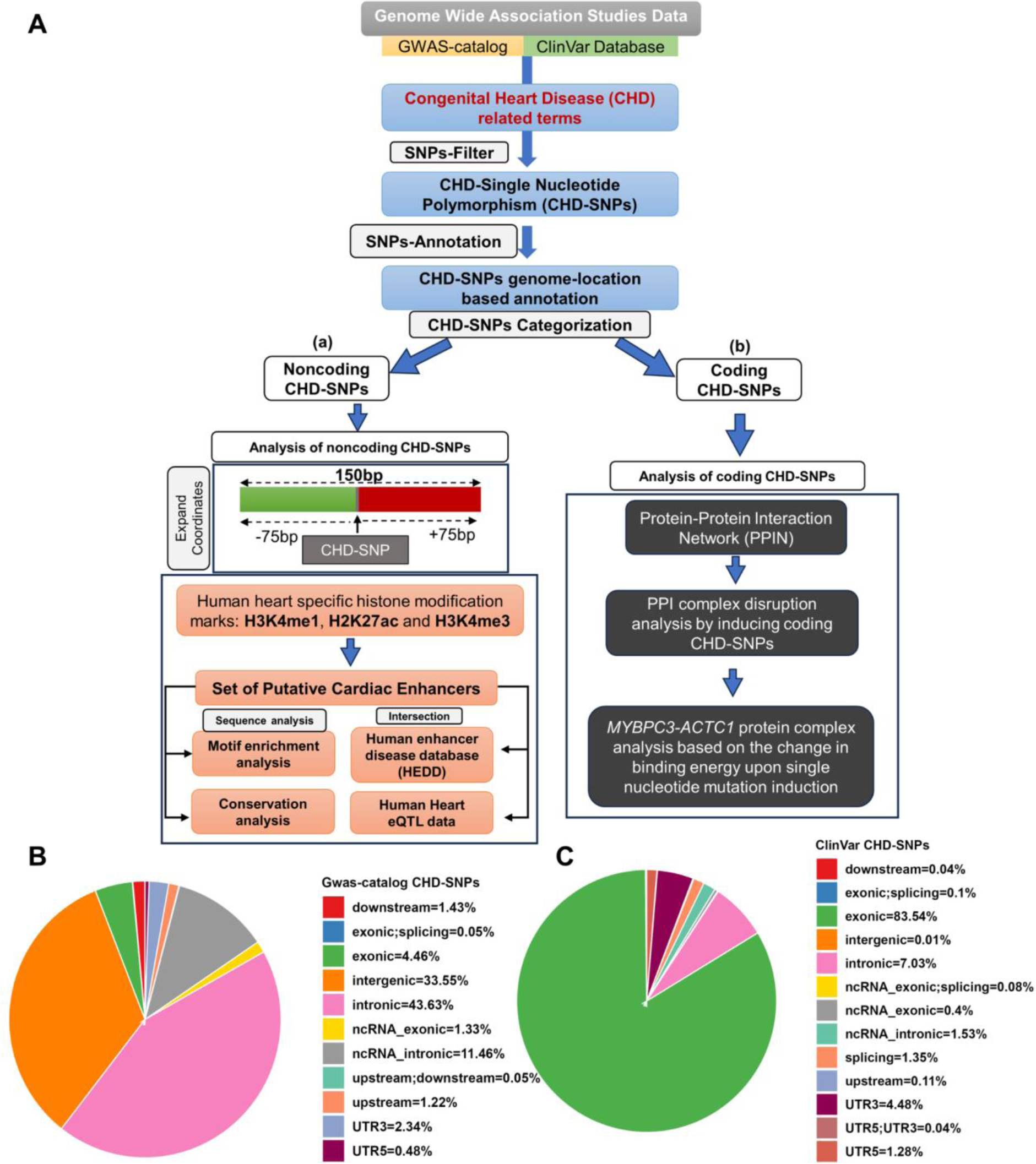
A computational workflow detailing the extraction of variant-trait data from genome-wide association studies, their filtration using congenital heart disease (CHD) associated terms and annotation using ANNOVAR. Post-filtration and annotation, CHD-SNPs undergo two-way analysis, depicted as (**a**) Analysis of noncoding CHD-SNPs and (**b**) Analysis of coding CHD-SNPs in the workflow. **B, C** Pie Charts illustrating the genomic distribution of CHD-SNPs obtained from GWAS-catalog and ClinVar database.

Based on their location, CHD-SNPs were classified into: (i) coding, indicative of sequences that transcribe into proteins, and (ii) noncoding, denoting sequences void of protein-coding potential. Noncoding regions specifically encompass distal intergenic, intronic, and promoter regions, the latter of which is demarcated by the stretch of region from 1kb upstream to 1kb downstream of the transcription start site (TSS). In total, out of 1,884 CHD-SNPs extracted from the GWAS-catalog, 163 and 1,721 were located in coding and noncoding regions of the genome, respectively. The latter consisted of 632 in distal intergenic, 822 in intronic, 216 in ncRNA-intronic and 51 in promoter regions. While in the ClinVar database, out of 13,392 CHD-SNPs, we obtained 12,171 within coding and 1,221 within noncoding genomic regions, with the latter consisting of 2 in distal intergenic, 984 in intronic, 214 in ncRNA-intronic and 21 in promoter regions.

### 2. Identification of a potential set of human cardiac enhancers

We then sought to ascertain the identity of the noncoding CHD-SNP containing regions as enhancers. Enhancer regions are enriched by specific histone modification marks which could also indicate their activity (27–30). Among these histone modification marks, H3K4me1 is associated with the presence of enhancers, while the presence of H3K27ac is a signature of active enhancers, differentiating them from their poised counterparts (31). Additionally, H3K4me3 predominantly marks promoter sequences (32,33). We hypothesize that noncoding CHD-SNPs might disrupt enhancer regions and consequently, TF binding. To test this, we first sought to assess the region carrying the CHD-SNP as functional enhancers. For this purpose, each noncoding CHD-SNP was expanded by 75bp on both flanks, yielding a 150bp genomic segment with the SNP centrally located. We then probed these segments for the presence of enhancer-specific histone modification marks. Utilizing epigenomic datasets (34) from human heart organogenesis across nine Carnegie stages (CS13, CS14, CS16-21, CS23), we sought regions marked by histone modification marks H3K4me1, H3K27ac, and H3K4me3 (pvalueSignal >= 9.0). Such elements exhibiting significant enrichment for these marks were designated as putative cardiac enhancers. Together, we obtained a set of 2,056 CHD-SNP-containing putative cardiac enhancers (CHD-enhancers) (**Table S4**), which consisted of 1,139 from GWAS-catalog and 917 from the ClinVar database. The analysis of CHD-enhancers distribution across Carnegie stages, illustrated in **Fig 2A**, unveils the dynamic landscape of CHD-enhancers shaping the embryonic development of the heart. Notably, the highest count of 1,138 enhancers identified in the later developmental stage (CS23) while in the initial stage (CS13) 821 enhancers were predicted. The lowest number of enhancers, 97, were observed in CS17. Moreover, the co-occurence of the predicted enhancers was also predicted, as shown in the intersection set in **Fig 2A**, where, for instance, out of the 821 identified enhancers in CS13, 530 were found to be expressed in other stages too, while 291 were specific to CS13. Furthermore, the occurrence of each predicted CHD-enhancer in a specific developmental stage, its presence in other stages, or its absence, revealed intricate patterns of regulatory activity throughout embryonic heart development (**Fig 2B**).

**Fig 2:**
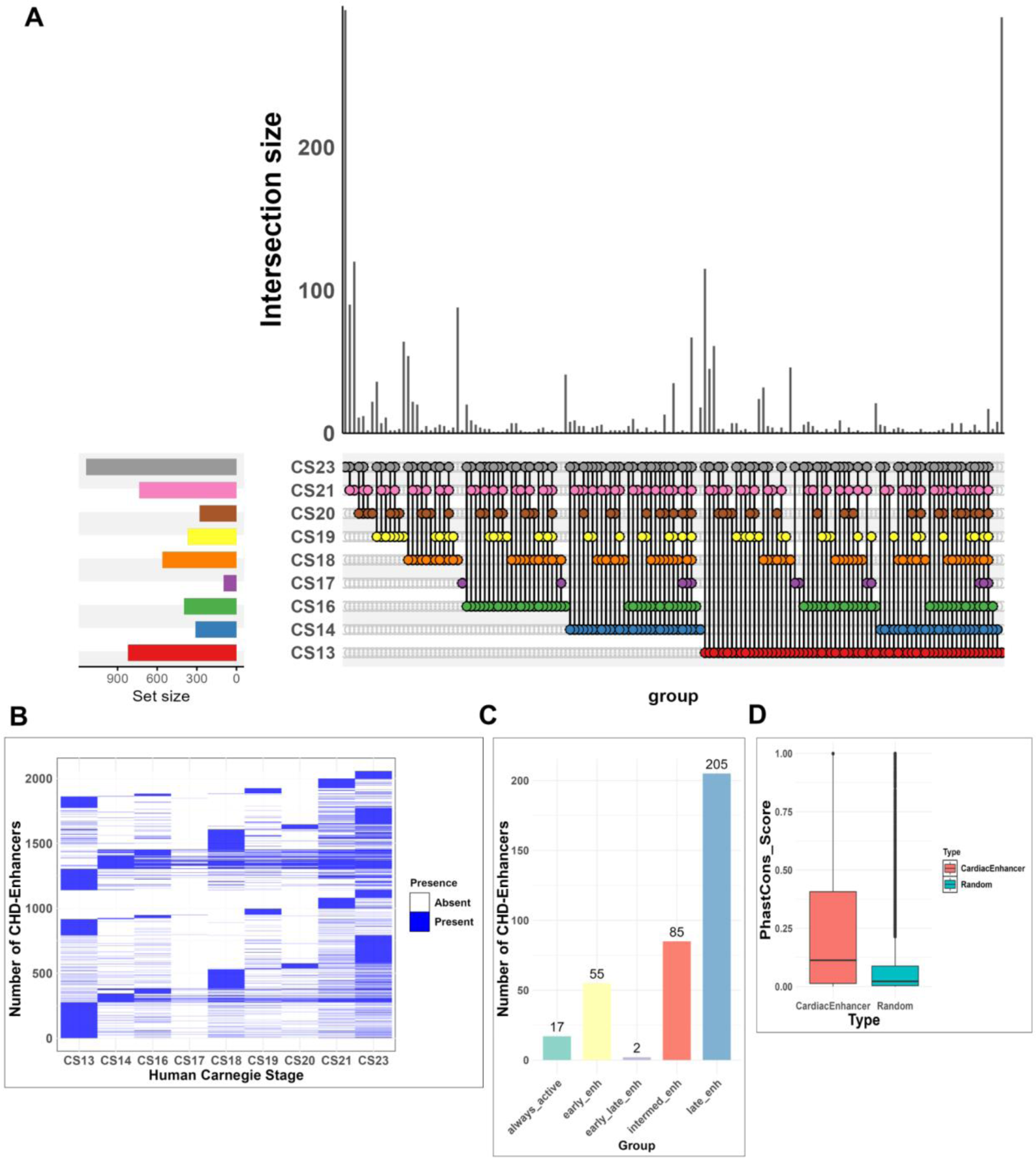
**A.** An upset plot showing the distribution and intersections of putative cardiac enhancers across nine Carnegie stages (CS13, CS14, CS15, CS16-21, CS23). Each vertical bar (shown in ‘*dark grey*’ color) indicates the number of occurrences of enhancers for an individual and combination of stages, while horizontal bars at the bottom denote the total enhancer counts for each stage. **B.** A heatmap representing the distribution of CHD-enhancers identified across nine distinct Carnegie stages (CS13, CS14, CS15, CS16-21, CS23). The color gradient ranging from ‘*white’* to ‘*blue*’ signifies the degree of enhancer-specific histone modification signals, where ‘*blue*’ marks the presence of a significant enhancer signal in a given stage and ‘*white’* indicates no detectable signal. **C.** A bar chart represents the stage-wise distribution of CHD-enhancers selected based on their presence in minimum 5 or more Carnegie stages. The categories include: (i) always_active enhancers: These show consistent activity across all Carnegie stages due to the presence of enhancer-specific histone modification marks. (ii) early_enh: represents enhancers active during Carnegie stages CS13,14,16. (iii) intermed_enh: active during stages CS17-19. (iv) late_enh: active during stages CS20,21,23. (v) early_late_enh: exhibits activity during both early and late Carnegie stages. **D.** A boxplot showing the comparison between conservation scores of putative cardiac enhancers (represented in ‘*salmon*’ colored box) and random noncoding genomic sequences (depicted in ‘*turquoise*’).

Subsequently, to corroborate our findings, we cross-referenced CHD-enhancers with human enhancer disease database (HEDD) (35). This intersection resulted in the identification of 801 enhancers that overlapped with CHD-enhancers (**Table S5**). Further, we categorized CHD-enhancers based on their *macs2pval* signal across all Carnegie stages. This resulted in 55 early, 85 intermediate, 205 late, 2 early-late and 17 always-active putative cardiac enhancers (**Fig 2C; Table S6**).

### 3. Enrichment of transcription factor binding motifs in putative cardiac enhancer regions

Enhancers regulate gene activity by binding to tissue-specific TFs at specific sites known as transcription factor binding sites or motifs (TFBSs) (36,37). Disease-associated noncoding variants within enhancer regions often perturb the TFBSs, resulting in disrupted transcriptional output of their target genes (38). To prioritize CHD-enhancers based on their potential regulatory function and study CHD-specific regulatory networks, CHD-enhancer regions were analyzed for the presence of TFBSs. From the motif enrichment analysis, we identified 163 statistically significant TFBSs (threshold on q-value < 0.05) (**Table S7**). Notably, many of these identified TFs play specific roles in early cardiac development, valvulogenesis, and cardiac maturation processes (**Table 1**). Among the TFBSs identified was that of Wilms’ tumor-1 (*Wt1*) (q-value=0.00023), known for its expression in cardiac endothelial cells during both normal heart development and post-infarction periods (39). Prior studies have elucidated its vital role in heart vessel formation (39). Another TF identified in this analysis was GATA Binding Protein 4 (*GATA4*) which is known to play a pivotal role in cardiac progenitor cells specification and cardiac septation (40). Additionally, our analysis revealed a significant over-representation of TFBSs from the Kruppel-like factor (*KLF*) family, recognized for their specialized roles in heart development (41). These included *KLF2* (42,43), *KLF4* (44,45), and *KLF15* (46), the latter shown to be overexpressed during episodes of heart pressure overload, typically seen in hypertrophic cardiomyopathies. It also regulates *GATA4* expression to stabilize cardiomyocyte size (46). Our analysis also identified the TFBS for Spleen focus forming virus proviral integration oncogene (*SPI1*), known for its heightened expression post-myocardial infarction (47). From the TEAD TF family, *TEAD1* was enriched, documented for its expression in adult mammalian hearts and association with normal heart contractility where its mutations can lead to dilated cardiomyopathy (48).

**Table 1:**
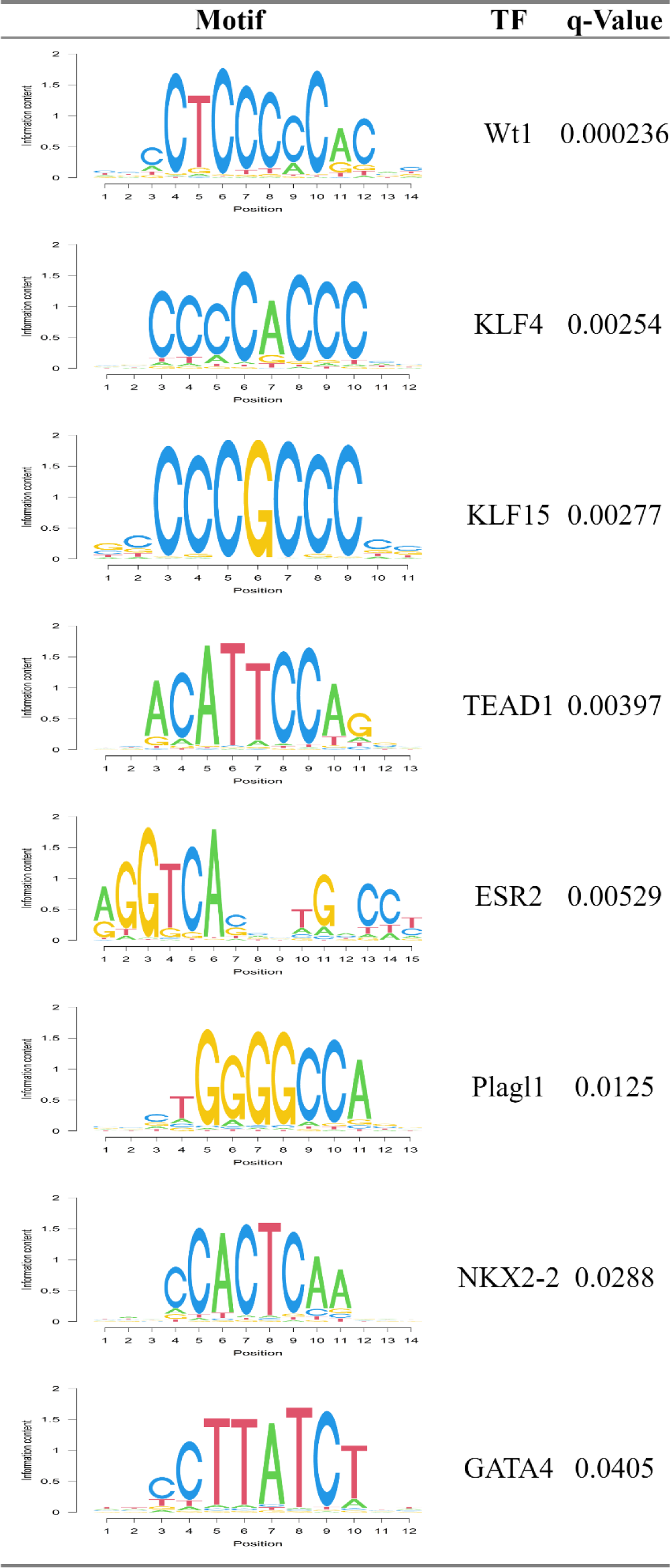
A list of significantly enriched Transcription Factor Binding Sites (TFBSs) identified within CHD-enhancers through motif enrichment analysis. The “Motif” column represents the DNA sequence of each binding site; “TF” specifies the transcription factor, and “q-Value” indicates the statistical significance level of the enriched motif.

| Motif | TF | q-Value |
| --- | --- | --- |
|  | Wt1 | 0.000236 |
|  | KLF4 | 0.00254 |
|  | KLF15 | 0.00277 |
|  | TEAD1 | 0.00397 |
|  | ESR2 | 0.00529 |
|  | Plagl1 | 0.0125 |
|  | NKX2-2 | 0.0288 |
|  | GATA4 | 0.0405 |

Other significant TFBSs enriched among the CHD-enhancers include Estrogen Related Receptor Alpha (*ESRRA*), known for its pivotal role in postnatal cardiac maturation through mitochondria-focused biogenesis in cardiomyocytes (49); PLAG1 Like Zinc Finger 1 (*PLAGL1*), important for sarcomere development and contractility in cardiomyocytes (50,51); and TGFB induced factor homeobox 1 (*TGIF1*), with an established role in a cardiac neural crest pathway crucial for the proper septation of the outflow tract (52). Together, the presence of these motifs and their linked TFs in our potential CHD-enhancers highlights their possible role in early heart development and maturation.

### 4. Putative cardiac enhancers and CHD-SNPs showed high conservancy across 99 vertebrate species

Although enhancers are known to be resistant to a certain degree of sequence mutations (53), it is noteworthy to highlight that enhancers with pivotal roles in developmental processes exhibit high conservation across vertebrates (54,55). We therefore sought to employ evolutionary conservation analysis of CHD-enhancers and their CHD-SNPs across 99 distantly related vertebrate species (56). We identified 131 conserved enhancers (131/2056, 6.37%) out of total 2,056 putative enhancers, with significantly high mean `phastCons score` ≥ 0.6 (55). Among these, 33 (33/2056, 1.60%) revealed ultra-high sequence conservation `phastCons score` ≥ 0.8 (57) (**Table S8**). To validate the significance of the evolutionary conservation observed in putative cardiac enhancers, we compared their conservation scores with randomly generated genomic sequences. This comparison allowed us to determine whether the observed conservation in our enhancers is indeed enhancer specific or merely reflective of broader genomic patterns. From this comparison, we obtained a mean conservation score of 0.2143 (*SD* = 0.2243) within the identified set of putative cardiac enhancers. In contrast, conservation analysis of random sequences revealed low mean conservation value 0.08264 (*SD* = 0.1501). Statistically significant differences were obtained between the conservation scores of compared groups with *p value* < 2.2*e*^−16^ from *Wilcoxon* rank sum test. The higher conservation scores for putative cardiac enhancers in comparison to random genomic elements suggest their vital evolutionary function (**Fig 2D**) and highlight their functional relevance in cardiac development. Furthermore, determining per-nucleotide conservation of CHD-SNP locations revealed 48 conserved variants with `phyloP score` ≥ 6.0 contained across 99 vertebrate species (**Table S9**).

To further determine the potential functional impact of CHD-SNPs on their target gene expression, we investigated the presence of regulatory SNPs (rSNPs) with known expression quantitative trait loci (eQTLs obtained from GTEx (58) database) in CHD-enhancers. We found 63 CHD-SNPs with known eQTLs which were distributed across various cardiac tissues, including 16 in the aorta, 9 in the coronary artery, 23 in the heart atrial appendage, and 15 in the left ventricle. These variants were designated as regulatory CHD-SNPs (rCHD-SNPs; **Table S10**). Notably, among rCHD-SNPs, an intronic variant *rs12724121* (chr1:236688982-236688983) was situated within a conserved CHD-enhancer element (chr1:236688907-236689057). This region aligns with DNaseI open chromatin regions from fetal heart tissue and also contained multiple TFBSs, including those specific to cardiac function (**Fig 3**). The presence of *rs12724121* in this evolutionarily conserved context suggests its potential regulatory role in cardiac-related processes. Moreover, the convergence of conservation, enhancer-specific histone modification marks, and TFBS density highlights the intricate regulatory landscape surrounding this particular CHD-SNP.

**Fig 3:**
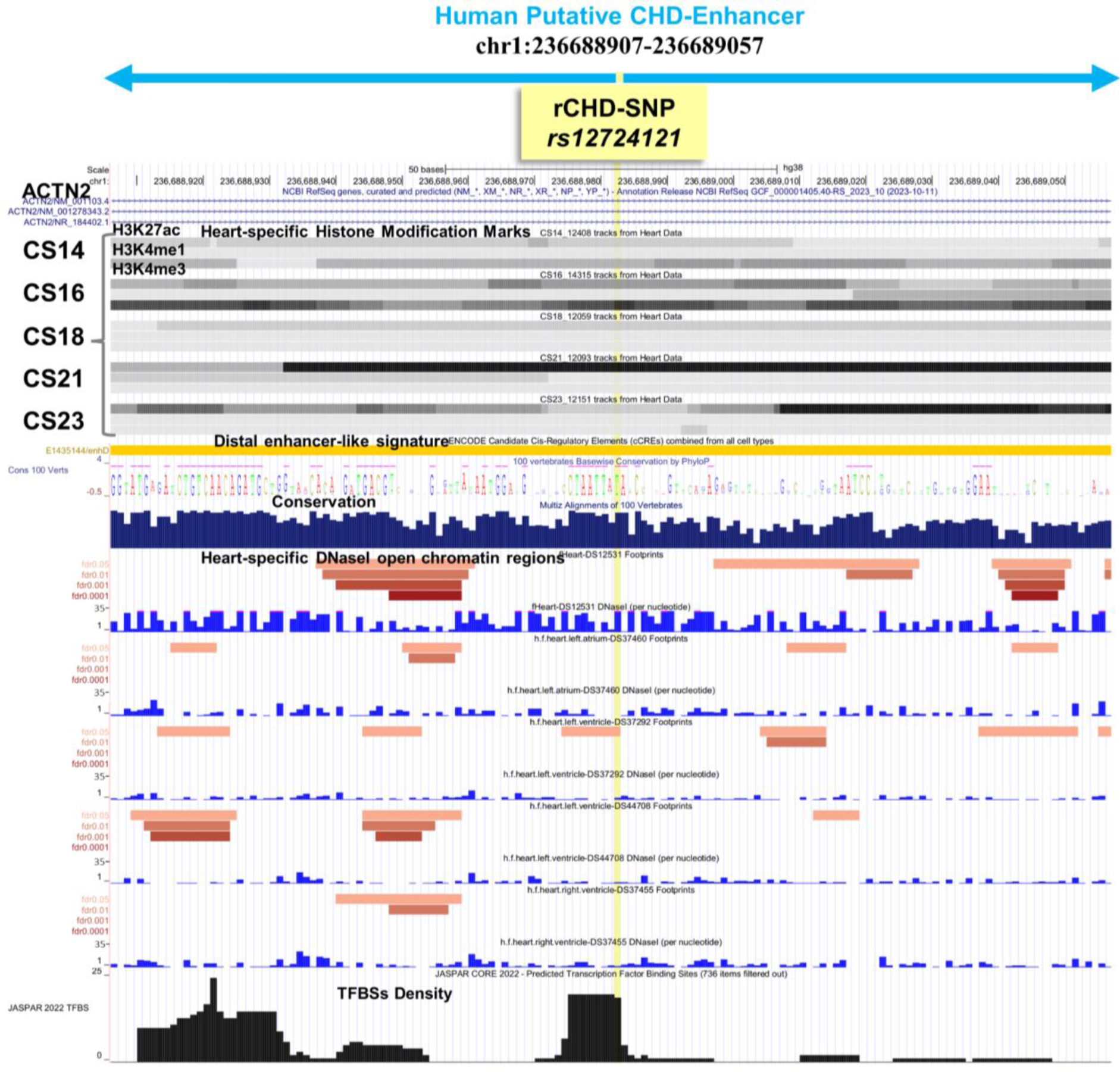
A screenshot captured from the UCSC genome browser illustrates a conserved human putative CHD-enhancer. The predicted enhancer resides within the intronic region of the *ACTN2* gene (chr1:236688907-236689057) and encompasses a regulatory CHD-SNP (rCHD-SNP; chr1:236688982-236688983 *rs12724121*, highlighted in *’yellow’*). The displayed enhancer region exhibits evolutionary conservation and is covered with heart-specific histone modification marks across 5 Carnegie stages (CS14, CS16, CS18, CS21, and CS23). It features a distal enhancer-like signature from the ENCODE registry of candidate *cis*-Regulatory Elements track. This region is further characterized by heart-specific DNaseI-hypersensitive sites (DNaseI), as obtained from the data by Vierstra *et al.* (81) and integrated as a UCSC track and exhibits overlap with a high density of transcription factor binding sites (TFBSs) from the Jaspar 2022 core vertebrates TFs database track.

### 5. CHD-SNPs within the coding regions can disrupt the protein-protein binding efficiency

SNPs within protein-coding genes can potentially disrupt protein-protein interactions, leading to abnormal cellular signaling and developmental defects that may contribute to CHD pathogenesis (17,59,60). To examine this, a protein-protein interaction network (PPIN) was constructed from the set of genes encompassing 11,770 coding CHD-SNPs. PPIN (**Fig S1**) consists of 306 proteins (286 genes from our query and 20 additional genes resulting from GeneMania-Cytoscape module output (61,62)) with 675 edges (connecting physically interacting proteins). The clustering analysis of the PPIN identified 16 functional modules. These clusters ranged in size, with the largest cluster, denoted as cluster 1 (**Fig S2**) encompassing 72 proteins and 304 edges. The smallest cluster consisted of only 3 nodes and 2 edges. Next, to study the impact of SNPs on the binding affinity of the physically interacting proteins, we selected two cardiac sarcomeric proteins from cluster-1 namely, cardiac myosin binding protein-C (*MYBPC3*) and cardiac α-actin (*ACTC1*) (16) (**Fig 4A, B**). Prior studies revealed a substantial number of individuals with hypertrophic cardiomyopathy (HCM) and dilated cardiomyopathy exhibit alterations in sarcomeric proteins (17,18). Therefore, to analyze the effect of CHD-SNPs on sarcomeric proteins, we obtained an experimentally resolved protein complex for *MYBPC3*-*ACTC1*(18) from protein data bank (PDB). Subsequently, the structural analysis of this protein complex (using PDBePISA (63)) revealed 15 interface residues and the remaining 331 residues were located on its surface. Changes in binding energy, a measure to assess protein complex stability (*ΔΔG*) was computed for 15 distinct mutations (exonic CHD-SNPs) and assessed their impact on protein binding stability using MutaBind2 (64) (**Table 2**). From this analysis, we found a specific CHD-SNP (*rs770030288*) which is located in C2 domain of *MYBPC3* protein with arginine to histidine substitution at 419 position has deleterious effect, which may impair its interaction with *ACTC1* protein. Notably, the evolutionary analysis of *MYBPC3* protein sequence using ConSurf (65) also detects high conservancy of arginine, highlighting the functional importance of this residue in preserving the conformational stability of the protein (**Fig 4C**). Moreover, upon inducing the R419H mutation, histidine (mutated residue) interacts with only 70 other atoms in vicinity (**Fig 4D (b, d)**) while the non-mutated form arginine interacts with 152 atoms, suggesting structural instability upon mutation (**Fig 4D (a, c)**).

**Fig 4:**
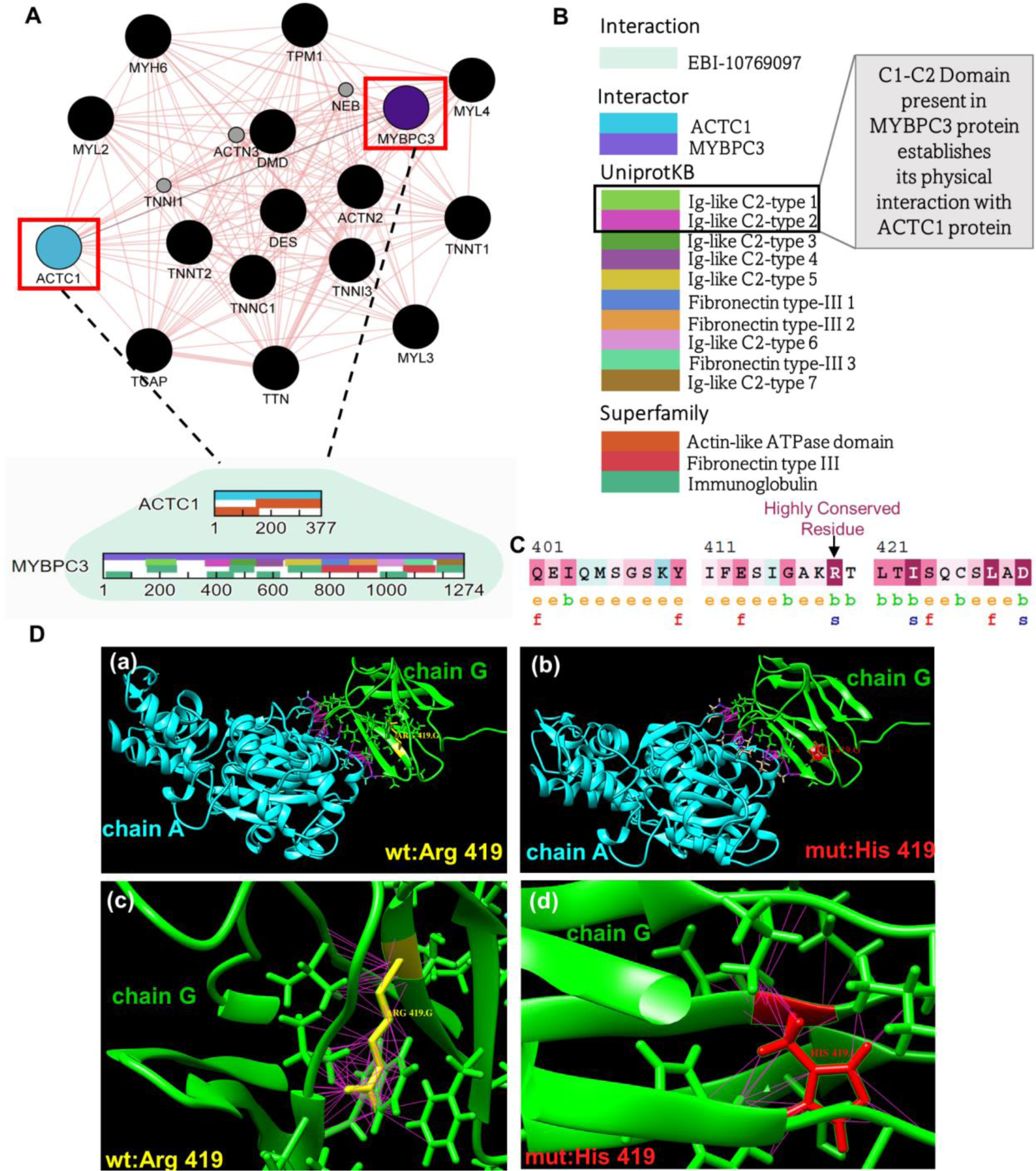
Analysis of the *MYBPC3*-*ACTC1* interaction in the PPIN. **A.** MCL-cluster1 from the PPIN (Shown in S1_Fig and S2_Fig) is illustrated with *nodes* representing proteins (‘*black*’ filled circles) and *edges* indicating direct physical interactions between them. Notably, the proteins *MYBPC3* and *ACTC1* are highlighted in ‘*purple*’ and ‘*blue*’, respectively, with their interaction denoted by a ‘*green*’ edge. **B.** Domain architecture in the interaction between *MYBPC3* and *ACTC1*. **C.** Evolutionary conservation analysis of *MYBPC3* protein showing conserved residues. **D. (a, b)** *MYBPC3:chainG* and *ACTC1:chainA* protein complex showing intermolecular contacts (‘*pink*’) within *ACTC1*:chainA (‘*cyan*’) and *MYBPC3*:chainG (‘*green*’) with wild-type residue ‘Arg’ (shown in ‘*yellow*’) and mutated residue ‘His’ (shown in ‘*red*’) at position 419 of *MYBPC3*:*chainG*. **(c, d)** Number of contacts (‘*pink*’) for wild-type residue ‘Arg419’ and mutated residue ‘His419’.

**Table 2:** Mutabind analysis of CHD-SNPs impact on Protein-Protein binding affinity. The columns “Protein 1 chain” and “Protein 2 chain” represents the protein 1 and protein 2 chains in a complex, “Mutated Chain” column represents the mutated protein chain, “Mutation” specifies the mutation, “DDG” quantifies the variation in free energy upon mutation and its impact on protein complex stability. A mutation is classified as deleterious if DDG ≥ 1.5 or ≤ −1.5 kcal mol^−1^. The column “Interface” indicates if the mutation occurs at the protein-protein interface or not.

| Protein 1 chain | Protein 2 chain | Mutated Chain | CHD-SNP | DDG | Interface | Deleterious |
| --- | --- | --- | --- | --- | --- | --- |
| A | G | G | **R419H** | **1.64** | no | **yes** |
| A | G | G | K380R | 0.44 | yes | no |
| A | G | G | E451Q | 0.03 | no | no |
| A | G | G | G416S | 1.35 | no | no |
| A | G | G | E441K | 1.23 | no | no |
| A | G | G | F448S | 0.29 | no | no |
| A | G | G | A364T | 0.41 | no | no |
| A | G | G | A429V | -0.02 | no | no |
| A | G | G | K380N | 1.08 | yes | no |
| A | G | G | K380R | 0.44 | yes | no |
| A | G | A | N92S | 0.3 | no | no |
| A | G | A | G245D | 1.27 | no | no |
| A | G | A | I267T | 0.6 | no | no |
| A | G | A | F21L | 0.44 | yes | no |
| A | G | A | L8M | 0.19 | no | no |

## Discussion

Our study aimed to investigate the contribution of SNPs located in both the noncoding and coding parts of the genome, and potential disruptions caused by them, on the gene regulatory system associated with congenital heart disease (CHD). Using publicly available human genomics data, we developed a computational pipeline to identify a robust set of SNPs associated with CHD. Significantly, a substantial portion of noncoding CHD-SNPs coincided with histone modification marks indicating enhancer activity specific for heart development. These regions, termed CHD-enhancers, were found enriched for important heart-specific transcription factor binding motifs, indicating their regulatory roles. Notably, a subset of identified CHD-enhancers, including the associated CHD-SNPs, exhibited a remarkable degree of conservation among vertebrates, implying their potential functional contribution in heart development. Moreover, in our analysis, a subset of the identified CHD-SNPs was discovered to align with known expression Quantitative Trait Loci (eQTLs). These specific genetic variants, termed regulatory CHD-SNPs (rCHD-SNPs), suggest a functional association with gene expression dysregulation.

From the coding CHD-SNPs, we studied how they might affect the protein-protein interactions in a network by introducing a series of mutations in protein structures. We predicted a highly conserved coding CHD-SNP (p. R419H) in cardiac sarcomeric protein *MYBPC3* which might negatively impact its interaction with another sarcomeric protein, *ACTC1*. A subset of CHD-associated SNPs in the noncoding region did not overlap with any enhancer marks. One possibility is that these SNPs could affect the function of noncoding transcripts. Many noncoding RNA have been implicated in heart development and disease (66,67). While our study focused on the effects of SNPs on enhancer region, those affecting the noncoding RNAs were not within the scope of our analysis. However, there are good examples of other studies dedicated to this topic. Zhu *et al.* presented a systematic methodology to analyze noncoding RNAs in the context of cardiovascular diseases which led to the identification of a novel ncRNA, *IGBP1P1*, whose depletion has been shown to restore cardiac function in disease (68). Liu *et al.* pinpointed key SNPs associated with Atrial Septal Defect as eQTLs for the lncRNA *STX18*-*AS1* (69). Another study by Hakansson *et al.* presented a link between SNPs within the 14q32 snoRNA locus and cardiovascular diseases in a substantial population-based investigation (70). As more and more noncoding transcripts are being identified their contribution to the CHD pathogenesis can be better understood.

With the increased capacity of genomics to discover disease-related SNPs, the repository will become increasingly enriched with *de novo* SNPs. Richter and colleagues (71) undertook such efforts, utilizing a machine learning approach to identify *de novo* noncoding variants by comparing whole genome sequences from 749 CHD patients with their parents and 1,611 child-parent healthy trios. Additionally, another study by Rummel *et al.* utilized massively parallel variant annotation to identify functionally important noncoding variants associated with Schizophrenia (72). These examples are expected to further expand our knowledge on the contribution of de novo mutations in various diseases and offer valuable insights into their mechanism. In conclusion, our study offers a systematic catalog of genetic variants associated with CHD. We explored their potential implications in impairing enhancer regulatory functions and disrupting protein-protein interactions. Our findings illuminate the intricate interplay between noncoding and coding SNPs, shedding light on a plausible mechanism contributing to CHD pathology.

## Materials and Methods

### 1. Data collection and computational framework design

A computational pipeline (**Fig 1A**) was designed to identify genetic variants associated with congenital heart disease (CHD) in both coding and noncoding regions of the genome. Data was collected from the GWAS-catalog (19) and ClinVar (20,21) database, capturing a broad range of genetic variations and their corresponding phenotypes, including both benign and pathogenic variants. From these datasets, CHD-specific SNPs were isolated based on 56 CHD-associated traits derived from several public databases and literature including “congenital heart disease”, “cardiac septal defects”, “heart structural deformities”, “non-syndromic CHD”, “CHD subtypes” (22–25) (**Table S1**). Next, ANNOVAR (26) was employed to classify these CHD-SNPs as either coding or noncoding, followed by specific computational analysis steps. For noncoding CHD-SNPs, genomic coordinates were expanded by 75bp both upstream and downstream of the SNP location, creating two subsets: one from the GWAS-catalog and another from ClinVar, each featuring a 150bp segment centered on the CHD-SNP (**Fig 1(a)**). These subsets then served in identifying putative cardiac enhancers (detailed in section 2 of methods). To study the impact of coding CHD-SNPs and determine the influence of missense mutations on the stability of protein complexes (detailed in sections 3 and 4 of methods), we constructed a protein-protein interaction network (PPIN) by incorporating genes containing CHD-SNPs in their exons (**Fig 1(b)**).

### 2. Identification of putative cardiac enhancers harbouring CHD-SNPs and their downstream analysis

Within extended genomic segments of 150bp, noncoding CHD-SNPs were probed for the presence of histone modification hallmarks indicative of enhancer activity, namely, H3K27ac, H3K4me1, and H3K4me3 (27–30). Notably, while H3K4me3 is predominantly linked with promoters, it can occasionally mark enhancers (32,33). For this analysis, epigenomic data from human heart organogenesis were obtained from Cotney’s lab (https://cotneylab.cam.uchc.edu/∼jcotney/HEART_HUB/hg38/primary/) (34). Intersections of these three histone modifications across nine Carnegie stages (CS13, CS14, CS16-21, CS23) with 150bp elements were computed. Subsequently, intersections with threshold of pvalSignal score >= 9.0 were retained. This step resulted in two sets of putative cardiac enhancers: one from the GWAS-catalog and the other derived from ClinVar database. Both sets were merged to a final unified set of putative cardiac enhancers which were further categorized based on their stage-wise developmental activity, including *early* (exhibiting high-pvalSignal in CS13, CS14 and CS16 for a given element of 150bp), *intermediate* (high-pvalSignal in CS17, CS18 and CS19 for a given element of 150bp), *late* (high-pvalSignal in CS20, CS21 and CS23 for a given element of 150bp) and *constitutively-active* (high-pvalSignal present in all stages for a given element of 150bp) elements. Additionally, pvalSignal was examined for stage combinations, encompassing early-intermediate, early-late, and intermediate-late. Following this, we investigated the presence of regulatory SNPs (rSNPs) within CHD-enhancers by utilizing cardiac eQTL data for Aorta, Coronary Artery, Heart Atrial Appendage, and Heart Left Ventricle from the GTEx database (58). Intersections between SNP locations in CHD-enhancers and eQTL SNP locations were performed and unique matches were retained.

In parallel, motif enrichment analysis was performed on the finalized set of putative cardiac enhancers using FIMO-MEME tool (73,74), with human genome as background. Transcription factor binding profiles were obtained from JASPAR Core database 2022 (75) (PWMs for vertebrates) to compute motif enrichments within enhancer sequences. The identified set of enriched motifs were filtered using threshold of q-value ≤ 0.05. Further, to determine evolutionary significance of identified enhancers, phastCons (55) conservation scores were computed from multiple sequence alignment across 99 vertebrate species downloaded from the UCSC genome browser (56). Average conservation signal within enhancer regions were computed using *bigWigAverageOverBed* (UCSC tools). Next, to determine the robustness of evolutionary conservation detected in CHD-enhancer elements, their conservation scores were compared to randomly generated noncoding genomic sequences. In this process, a set of 10,000 random sequences of 150bp were generated from noncoding human genome and their average conservation signal (*phastCons*) was computed across 99 vertebrate species. Mean conservation scores were compared between putative enhancer and random genomic sequences and statistical significance was computed using *wilcoxon* rank sum test. Further, per-base conservation for CHD-SNPs locations was also determined by calculating *phyloP* (57) conservation scores across 99 vertebrate species. As validation, CHD-enhancer coordinates were cross-referenced with known enhancers from human enhancer disease database (35).

### 3. Protein-Protein interaction network construction from coding CHD-SNPs and protein sequence analysis

To analyze a set of coding CHD-SNPs, PPIN was constructed using GeneMania (61), a plugin within the Cytoscape tool (62). In this network, ‘nodes’ represented genes containing exonic CHD-SNPs, and the ‘edges’ illustrated the interactions between physically interacting proteins. Subsequently, clusters within the PPIN were identified based on markov cluster algorithm (76) (MCL; **Fig S2**) from *clusterMaker* plugin (77). To discern functionally significant regions within protein sequences, protein domain analysis was conducted using the Simple Modular Architecture Research Tool (78) (SMART) together with evolutionary conservation analysis using conSurf tool (65). Additionally, for the detection of domains responsible for establishingprotein-prtoein interactions IntAct molecular interaction database was utilized (https://www.ebi.ac.uk/intact/home).

### 4. Coding CHD-SNPs and their impact on protein-protein interactions

The impact of missense mutations (nonsynonymous coding CHD-SNPs) on protein-protein complex stability (59,60) was evaluated by obtaining a three-dimensional structure of a protein-protein complex from the protein data bank (79). Single mutations were introduced within interacting monomers of the complex using structure editing (rotamers) tool of Chimera (http://www.cgl.ucsf.edu/chimera/) (80). Further, change in binding free energy (ΔΔG_bind_) was computed using MutaBind2 (64) for each of the mutated complex, providing a quantitative measure of the mutational impact on protein–protein interaction stability.

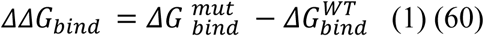

Binding energy *ΔG*_*bind*_ is computed by taking the difference between free energies of bound proteins (AB) and protein monomers in unbound state (A and B):

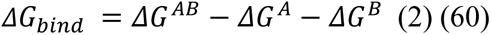

## Acknowledgements

This work was supported by the Polish National Science Center (NCN) OPUS grant 2018/29/B/NZ2/01010 and 2022/47/B/NZ2/02926. We thank all the members of ZDG lab for fruitful discussions. We would like to thank M. Pawlak for initial support in the project.

## Supplementary Information

The supplementary information includes Excel files containing supplementary tables and a document with supplementary figures.

## Data Availability Statement

The source code and data used to produce the results and analyses presented in this manuscript are available at https://github.com/rshikha95/CHD-SNPs.git.

