## Supplementary figures and images for "Computational analysis of congenital heart disease associated SNPs: Unveiling their impact on the gene regulatory system"

### Supplemental Figure 1

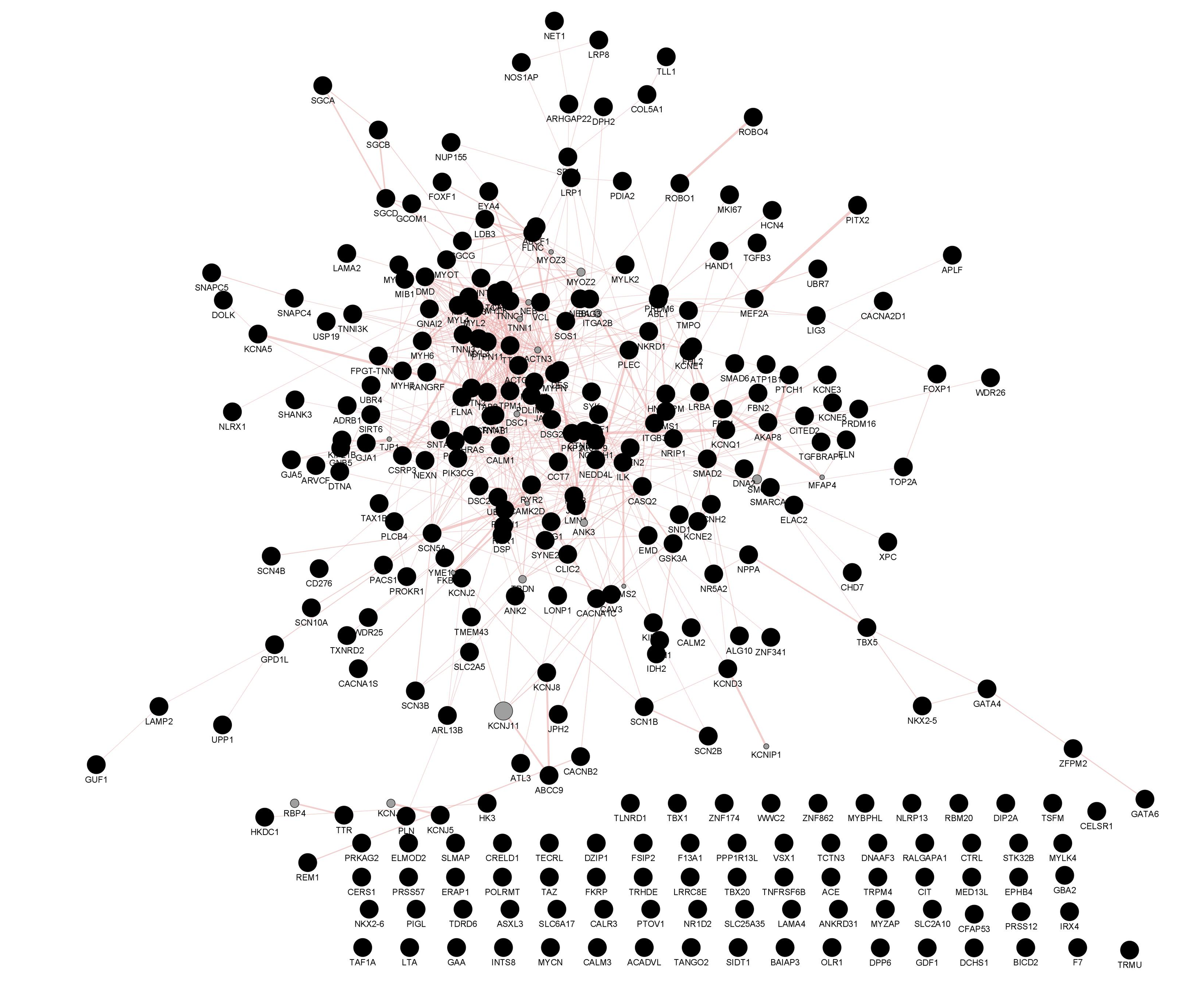

### Supplemental Figure 2

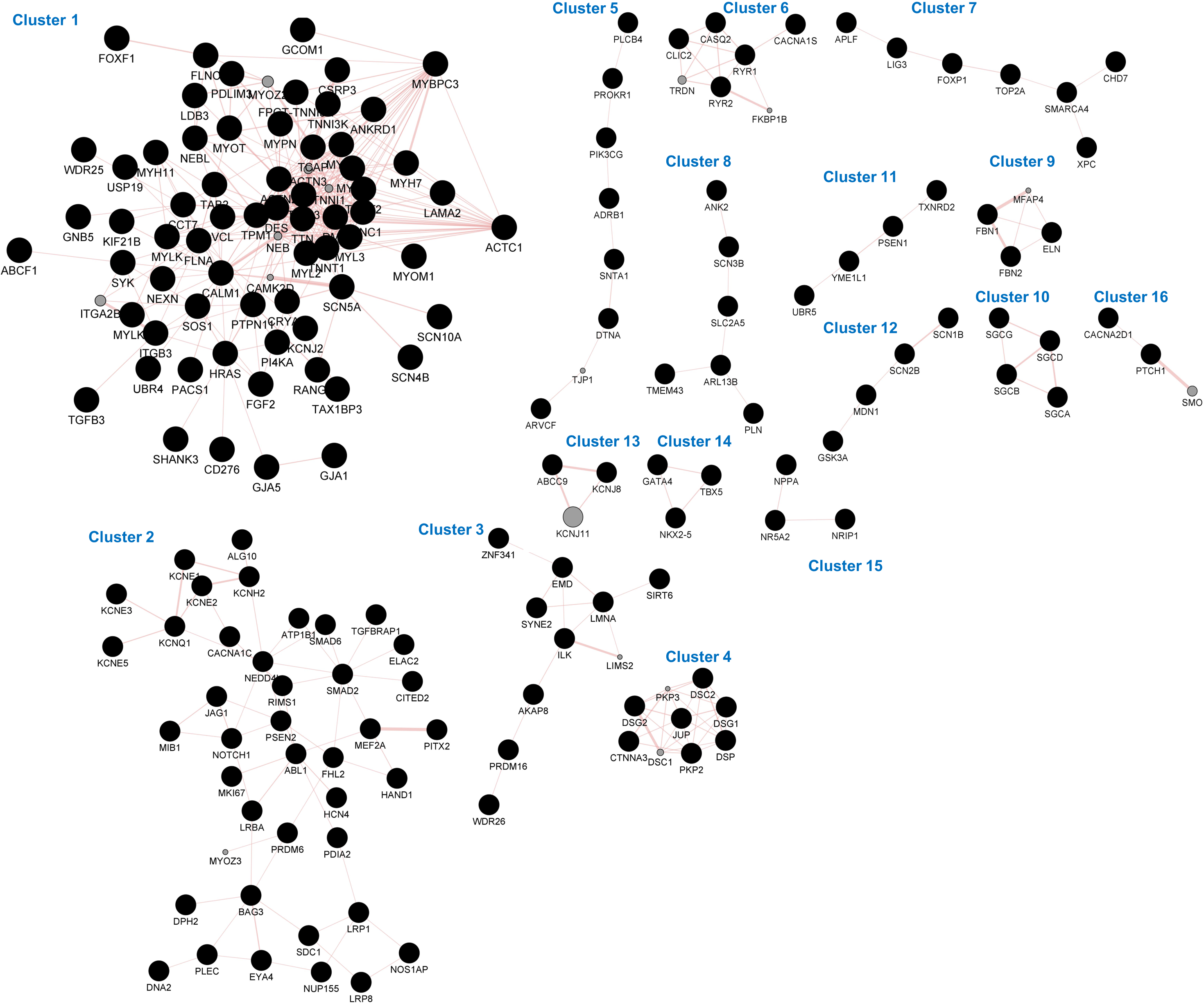
