## Supplemental Information for "Computational analysis of congenital heart disease associated SNPs: Unveiling their impact on the gene regulatory system"

### A computational approach to study SNPs associated with congenital heart disease

#### Supplementary Figures

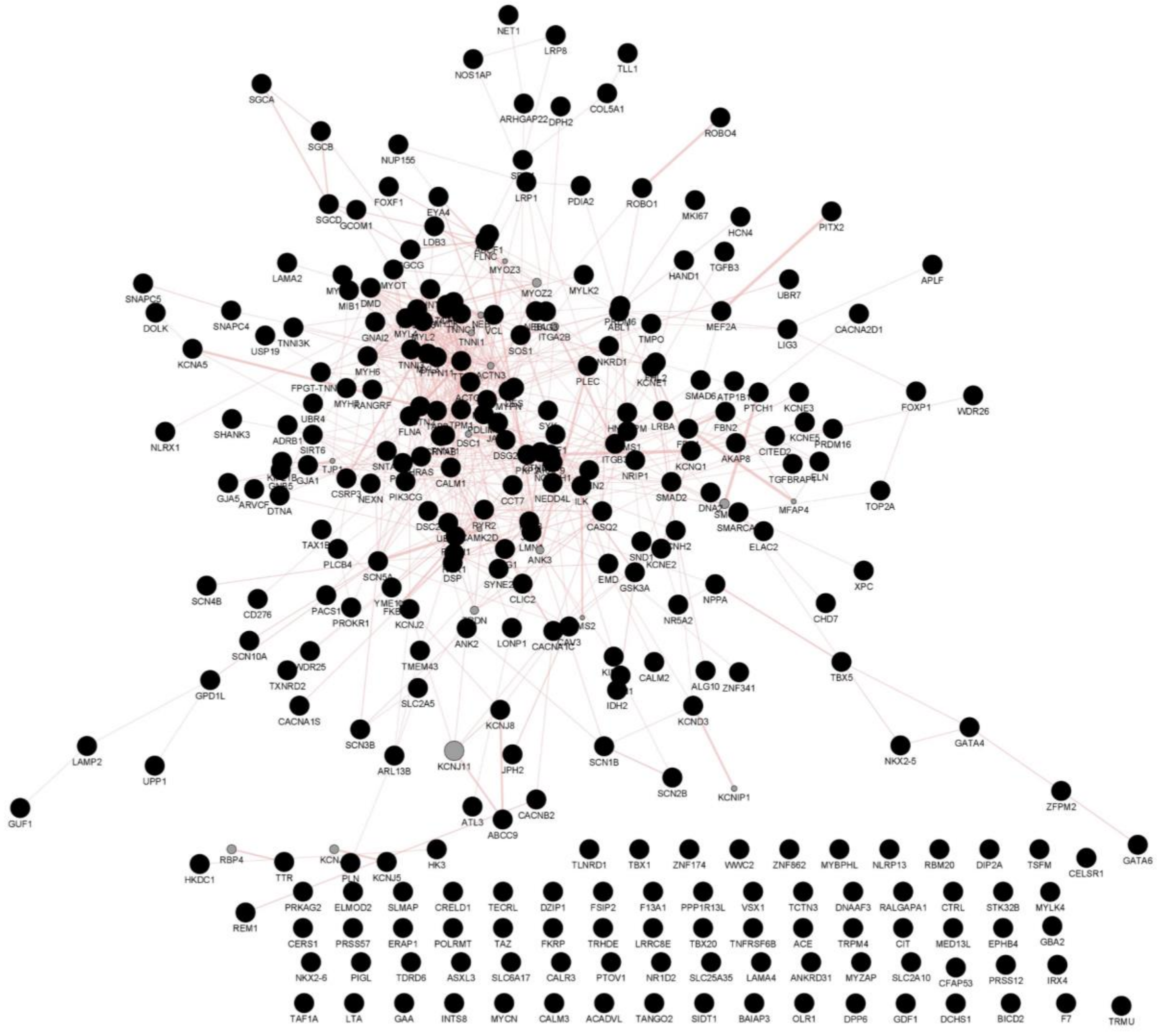


**S1_Fig.** Protein-protein interaction network (PPIN). In the PPIN, *nodes* represent proteins containing coding CHD-SNPs, and the *edges* signify interactions between physically interacting proteins. The ‘*black’* nodes correspond to query proteins and ‘*grey’* nodes represent additional proteins in the network identified in the GeneMania query search. The size of each *'grey'* node is determined by the gene score, reflecting the relatedness of these proteins according to GeneMANIA. Larger circles indicate a stronger predicted connection between these proteins. Width of the edges signifies the weight of interaction between proteins. Thicker edges indicate stronger physical interactions. The network is constructed using the cytoscape-GeneMANIA module.

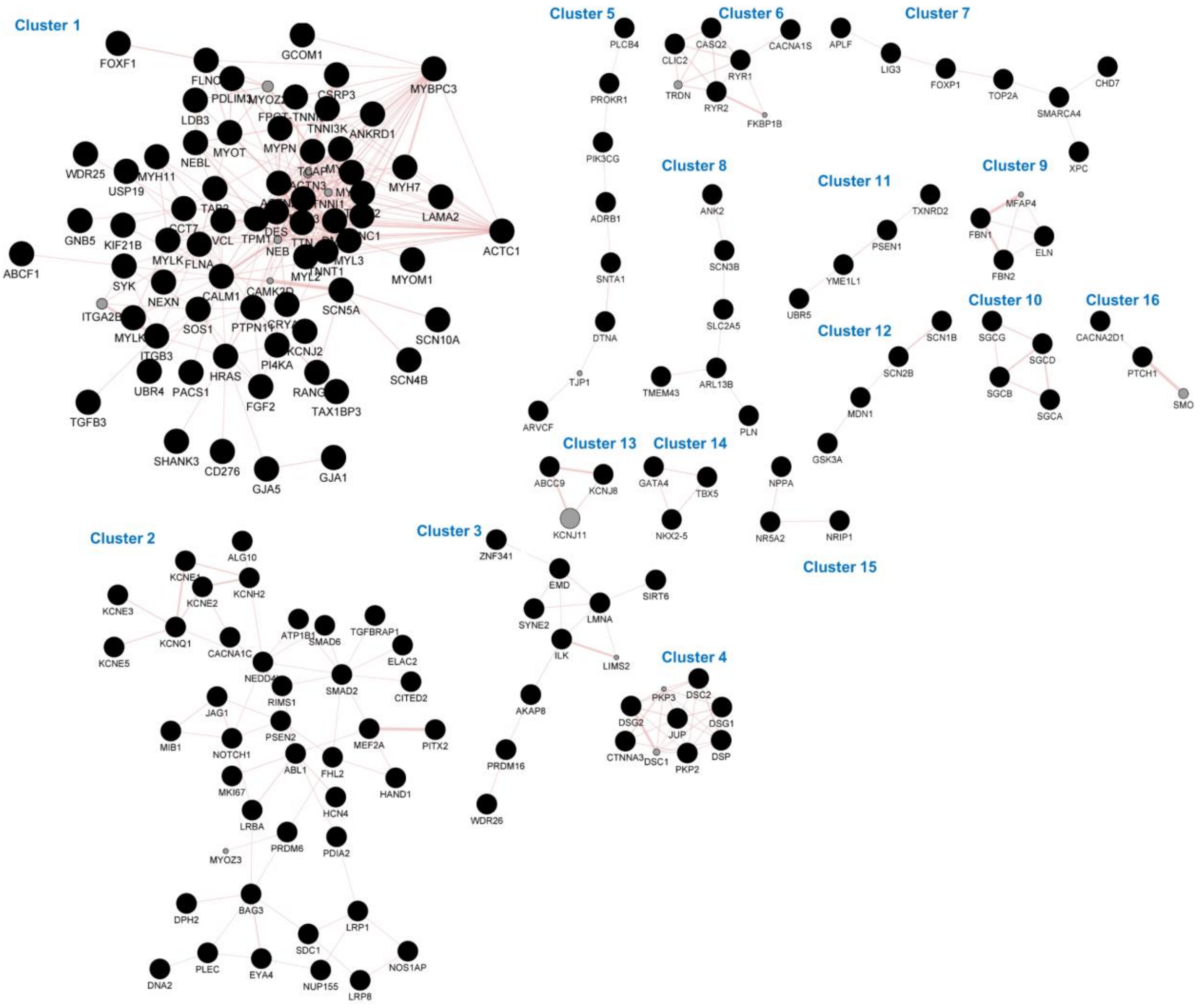


**S2_Fig:** Clustering of protein-protein interaction network using MCL algorithm.

The figure comprises 16 distinct clusters numbered from 1 through 16. In this network, *nodes* represent proteins containing coding CHD-SNPs, and the *edges* signify interactions between physically interacting proteins.
